# Resting-state EEG oscillations are reduced in asymptomatic *C9orf72* repeat expansion carriers

**DOI:** 10.1101/2025.08.19.25333576

**Authors:** Stefan Dukic, Kevin van Veenhuijzen, Annebelle Michielsen, Henk-Jan Westeneng, Leonard H. van den Berg

**Affiliations:** Department of Neurology, University Medical Center Utrecht Brain Center, Utrecht University, Utrecht, The Netherlands

## Abstract

**Objective:** We investigated the signature of the *C9orf72* repeat expansion on resting-state cortical activity in asymptomatic individuals and explored its relationship with structural, gene expression, and cognitive measures.

**Methods:** High-density resting-state EEG was compared between 90 asymptomatic family members of patients with familial amyotrophic lateral sclerosis (ALS), dichotomised into carriers of the pathological *C9orf72* repeat expansion (N = 37) and non-carrier controls (N = 53). Periodic (oscillatory) and aperiodic (1/f) components of the power spectrum were analysed at the sensor- and source-level. Regional EEG power changes were correlated with *C9orf72* expression from the Allen Human Brain Atlas, cortical MRI thickness and performance on the Complementary Cognitive ALS Screen (C-CAS).

**Results:** Asymptomatic carriers exhibited significantly lower periodic power in the α and β frequency bands (10-30 Hz) across posterior cortical regions, including the parietal, occipital, and temporal lobes. The magnitude of this power reduction was associated with increased *C9orf72* expression and reduced cortical thickness in the same regions. In carriers, reduced β-band power was associated with poorer performance on Visuoconstructive Immediate and Body Representation tasks. The aperiodic component of the EEG power did not differ between groups.

**Interpretation:** In asymptomatic *C9orf72* repeat expansion carriers, resting-state EEG reveals differences in oscillatory power in the posterior brain regions. The correlational findings suggest that *C9orf72* repeat expansion may be involved in functional and structural changes in the posterior cerebral cortex, which may contribute to deficits in tasks requiring visuospatial processing.

## Introduction

Early diagnosis in amyotrophic lateral sclerosis (ALS) remains a major clinical challenge, as therapeutic outcomes are likely dependent on intervention before irreversible neuronal damage occurs. While the majority of cases are sporadic, a significant subset is linked to a GGGGCC hexanucleotide repeat expansion (RE) in the *C9orf72* gene,^1^ a mutation with incomplete penetrance^2^ and considerable clinical overlap with frontotemporal dementia (FTD).^3^ Understanding the direct impact of this mutation on brain function during the asymptomatic phase is, therefore, crucial for identifying sensitive, early biomarkers and elucidating the initial stages of brain dysfunction.

Previous research in asymptomatic *C9orf72* RE carriers has consistently identified structural brain changes using MRI,^4^ with our own recent work demonstrating grey matter alterations not only in the motor cortex and thalamus, but also in parietal, occipital, and temporal regions.^5^ We have also recently provided functional evidence from task-based EEG, showing increased engagement of parietal areas during a sustained attention task in an overlapping cohort.^6^ To date, however, the functional integrity of brain regions activated under different conditions remains to be elucidated. The resting-state paradigm offers a significant advantage over task-based approaches by circumventing potential confounds such as administrative difficulty and variations in participant motivation, making them highly suitable for biomarker development.

To address this gap, the present study aimed to evaluate resting-state cortical regions in asymptomatic *C9orf72* RE carriers using high-density EEG. We characterised changes in EEG power and investigated their association with structural, gene expression, and cognitive data. To isolate the effects of the *C9orf72* RE while minimising confounding genetic and environmental factors, our cohort was comprised exclusively of related asymptomatic individuals from families with a known history of ALS.

## Methods

### Participants

Between December 2020 and June 2025, a cohort of 100 family members of patients with *C9orf72*-associated familial ALS were recruited. All participants were recruited through relatives diagnosed with ALS at the Motor Neuron Disease Outpatient Clinic of the University Medical Centre Utrecht. All participants were 18 years of age or older and provided written informed consent in accordance with the Declaration of Helsinki. This study was approved by the medical ethics committee of the University Medical Centre Utrecht.

From this initial cohort, 10 participants were excluded due to: missing cognitive or neurological examination results (N = 3); abnormal cognitive test outcomes (N = 3); or confounding factors such as the use of psychoactive medication or a history of significant head trauma (N = 4). The remaining 90 participants were classified as asymptomatic family members (AFM), defined by the absence of clinical signs of motor neuron disease, bulbar dysfunction, or cognitive impairment, as previously described.^5,6^ This included a standardised neurological examination to rule out signs of upper or lower motor neuron involvement^7^ and cognitive testing using the Dutch version of the Edinburgh Cognitive and Behavioural ALS Screen (ECAS) to exclude cognitive symptoms.^8^

Genetic testing for the *C9orf72* RE was performed on genomic DNA samples, with pathological expansions defined as ≥30 GGGGCC repeats.^9^ Based on this criterion, the final cohort was dichotomised into AFM with the pathological *C9orf72* RE (C9+) and those without (C9−).

### Experimental paradigm

The experimental paradigm was resting-state with eyes closed. The session was divided into three 2-minute blocks, separated by short breaks of less than one minute. Participants were seated comfortably and asked to relax, minimise eye movements, and let their minds wander.

### EEG data

High-density EEG data were acquired using a 128-channel BioSemi Active Two system (BioSemi B.V., Amsterdam, The Netherlands). The signal was sampled at 512 Hz and low-pass filtered at 104 Hz by the acquisition hardware. Preprocessing was conducted using custom automated MATLAB scripts, as previously described.^6^ Cleaned signals were segmented into 2-second epochs with a 50% overlap, re-referenced to the common average, and baseline-corrected by subtracting the mean amplitude. Epochs were then automatically screened for artifacts, and remaining epochs were used for subsequent analysis. The breakdown of preprocessing outcomes and data quality assessments is detailed in Supplementary Table 1.

Source localisation was performed using the exact Low Resolution Electromagnetic Tomography (eLORETA)^10^. Individualised, realistically shaped boundary element models were constructed based on each participant’s T1-weighted MRI scan, while for five participants lacking an MRI scan a model based on the ICBM152 template^11,12^ was used. The models incorporated isotropic conductivity values for brain (0.33 S/m), skull (0.008 S/m), and scalp (0.43 S/m).

A source model of the cortex was estimated with 164,000 vertices per hemisphere and subsequently downsampled to 8,000 vertices per hemisphere using FreeSurfer.^13^ For leadfield whitening, a diagonal matrix containing the average power of EEG signals, high-pass filtered above 50 Hz to approximate noise from muscle activity, was used for each participant. The signal-to-noise ratio, required for regularisation in eLORETA, was set to three. Source signals were then estimated for 68 cortical regions defined by the Desikan-Killiany (DK) atlas.^14^ For each participant and brain region, an equivalent signal was estimated by first extracting a signal oriented perpendicular to each vertex, and then applying singular value decomposition to the DK region’s source-space covariance matrix to extract the strongest component.

Power spectral density of each epoch was calculated using Welch’s function in MATLAB, employing a 2-second Hamming window. The resulting power spectra were then parameterised to separate the periodic (oscillatory) and aperiodic (1/f) components using the Spectral Parametrisation (specparam) method, formerly known as Fitting Oscillations and One Over *f* (FOOOF).^15^ The model was fitted across a 2–48 Hz frequency range with the aperiodic mode set to “fixed”, a maximum of four peaks, peak width limits of 1–8 Hz, minimum peak height of zero, and all other parameters left at their default settings. Sensitivity analyses with varied parameters primarily concerning the frequency range confirmed the robustness of the primary results (data not shown).

For subsequent analyses, the aperiodic component, consisting of the offset and slope, was taken directly from the fitted model. The periodic component was calculated as the residual power spectrum after subtracting the aperiodic fit from the original raw spectrum. This approach was chosen to circumvent occasional poor model fitting of oscillatory peaks sometimes observed in the lower frequency range. From this periodic component, total power of each frequency band (N = 7) was calculated: *δ* (2–4 Hz), *θ* (4–8 Hz), *α* (*α*_l_: 8–10 Hz, *α*_h_: 10–13 Hz), *β* (*β*_l_: 13–20 Hz, *β*_h_: 20–30 Hz), and *γ* (30–48 Hz).

To assess the excitation/inhibition balance, an aperiodic slope, a proposed proxy for this physiological state, was quantified in each electrode.^16^ As there is no consensus on the optimal frequency range for this estimation, and given that the broadband range (2–48 Hz) is not typically used for this purpose, aperiodic slopes in the commonly used higher-frequency range of 30–80 Hz were estimated as well.

### MRI, gene expression and cognitive data

A 3T Philips (Best, the Netherlands) Achieva Medical Scanner was used to acquire T1-weighted images (N = 85), with MRI parameters and preprocessing as described previously.^17^

Cortical *C9orf72* expression data were obtained from the Allen Human Brain Atlas (AHBA), a public dataset comprising postmortem microarray data from six adult donors (1 female, mean age = 42.5 ± 13.4 years) with no known neuropathological conditions.^18^ Processing of these data was performed as previously described,^5^ with the exception that a min-max scaling transformation was applied to the unit interval in order to facilitate interpretability. As expression data were available from all six donors for the left hemisphere but only two for the right, all subsequent analyses were restricted to the left hemisphere to ensure consistency.

Scores from the Complementary Cognitive ALS Screen (C-CAS), a screening tool designed to assess cognitive domains beyond those covered by the ECAS, such as functions associated with the parietal cortex.^19^ Raw scores (N = 65) were converted to residuals using normative models that accounted for age, sex, and education when necessary, with higher residual scores indicating poorer performance.

### Statistical analysis

Differences in demographics between groups were assessed using the Mann–Whitney U test for continuous variables and Fisher’s exact test for categorical variables.

Neuropsychological performance on ECAS was compared using linear regression, with ECAS scores as the response variable and age, sex, education level (high/low), and *C9orf72* RE carriership as predictors. For C-CAS, neuropsychological performance was compared using the Mann–Whitney U test using the obtained score residuals.

Upper motor neuron function was compared using binomial logistic regression for dichotomous outcomes or ordinal regression for outcomes with more than two categories. For these analyses, the assessment outcome served as the response variable, with age, sex, and *C9orf72* RE carriership as predictors. For each assessment outcome, the most deviant neurological sign, irrespective of laterality, was used in the analysis.

Group differences in EEG measures were evaluated using a linear mixed-effects model, which included age, sex, and *C9orf72* RE carriership as fixed effects. To account for dependencies within the EEG data and familial relationships, a random intercept for pedigree ID was added. Prior to this analysis, EEG measures were transformed to standard normal distributions using the inverse normal transformation due to their non-normal distribution,^20^ and separate linear models were fitted for each electrode and brain region. Multiple comparisons were controlled using the threshold-free cluster enhancement method (E = 0.5, H = 2)^21^ with 5,000 permutations, using a neighbourhood radius of 40 mm. Statistical significance was set at P < 0.05, corrected for multiple comparisons.

A sensitivity analysis was conducted to ensure the robustness of the primary findings against two potential confounds: a marginal age difference between groups (P = 0.05) and the inclusion of single-participant pedigrees (N = 16). To address both factors simultaneously, the primary statistical analysis was repeated on a reduced cohort (N = 64) that excluded C9− AFM under the age of 30 and all individuals from single-participant pedigrees. This adjustment resulted in more comparable age distributions between groups (P = 0.84) without introducing other demographic confounders. As this sensitivity analysis yielded the same pattern of group differences as the primary analysis (data not shown), the results from the complete dataset (N = 90) are presented in the main text.

To assess the relationship between the spatial patterns of EEG power differences and gene expression, a spatial autocorrelation-preserving permutation test ("spin test")^22^ was conducted. First, a surface-based representation of the DK atlas was created on the FreeSurfer fsaverage surface. The spatial coordinates for each cortical region were defined as the vertex closest to the region’s centre of mass. To generate a null distribution, these coordinates were randomly rotated on the spherical projection of the surface, and the original regions were reassigned the expression values of the spatially closest rotated region.^23^ For each permutation (N = 10,000), a meta-regression analysis was performed across all cortical regions, using the regional *C9orf72* expression data from the AHBA as a moderator for the group differences in periodic EEG power (represented by the β-coefficients from the primary linear mixed-effects models).

To assess the relationship between the spatial patterns of EEG power and structural differences, a correlation analysis was conducted. Spearman’s rank correlation was used to relate regional group differences in EEG power with the regional group differences in cortical thickness across both hemispheres, with both measures represented by their respective β-coefficients from liner mixed-effects models with the same structure as in the primary analysis.

To determine the functional relevance of our findings, linear mixed-effects models were used to test whether the relationship between EEG power and cognitive performance on C-CAS was moderated by *C9orf72* RE carriership. The dependent variable in this model was the EEG power, estimated as an average across the regions significantly different between AFM C9- and AFM C9+ identified in the main group analysis and weighted by their respective P-values. Besides age and sex, independent variables were the C-CAS residuals, *C9orf72* RE carriership, and their interaction term.

## Results

### The demographic profiles

In total, 90 AFM were analysed of which 53 C9− and 37 C9+. Total number of pedigrees was 37, with a median number of AFM per pedigree: 2 (range: 1-15). All participants were up to the third degree of consanguinity with respect to the closest family member with ALS. Demographics of the participants are summarised in Table 1.

**Table 1.**
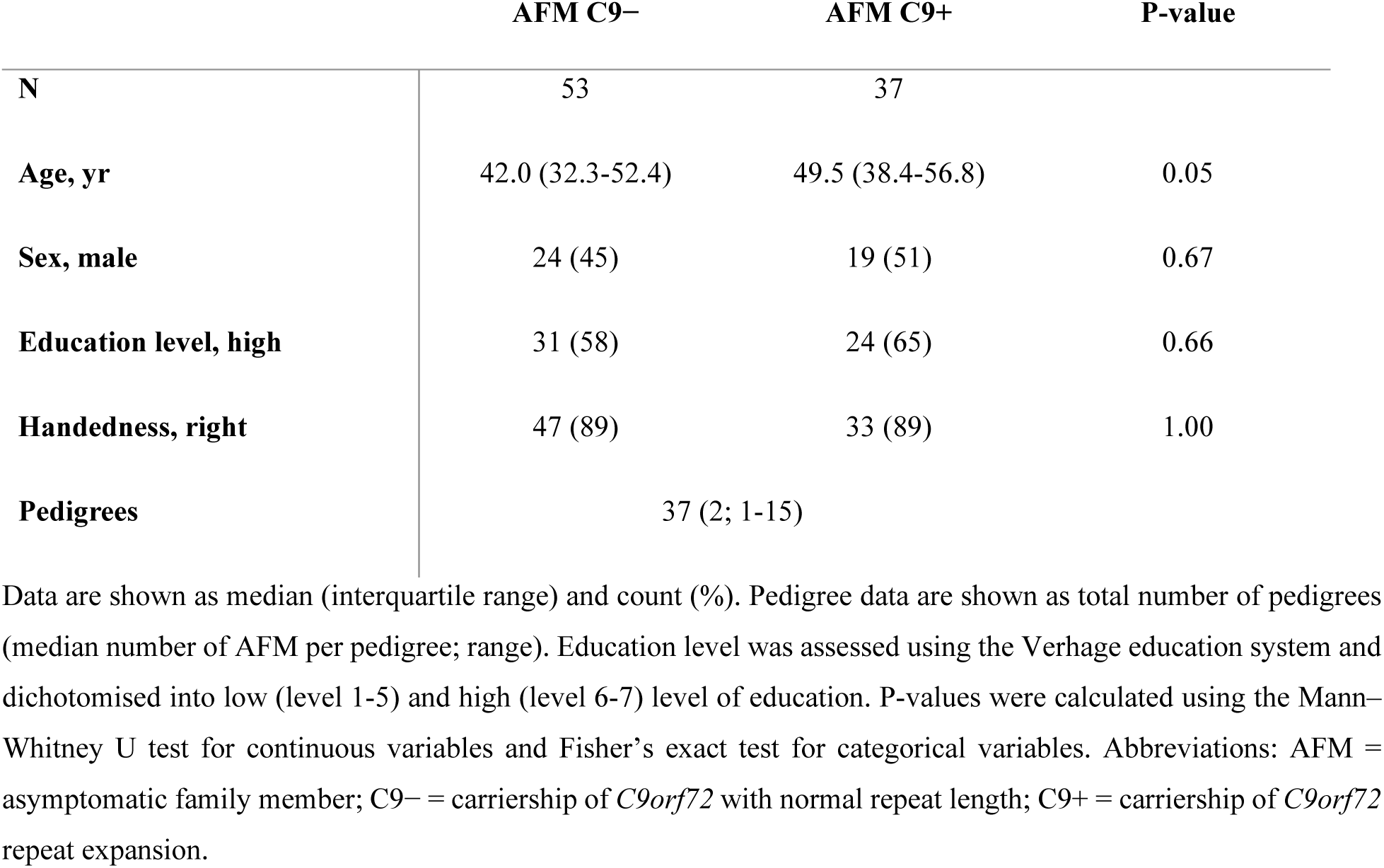
Demographic and clinical characteristics of asymptomatic family members.

|  | AFM C9– | AFM C9+ | P-value |
| --- | --- | --- | --- |
| N | 53 | 37 |  |
| Age, yr | 42.0 (32.3-52.4) | 49.5 (38.4-56.8) | 0.05 |
| Sex, male | 24 (45) | 19 (51) | 0.67 |
| Education level, high | 31 (58) | 24 (65) | 0.66 |
| Handedness, right | 47 (89) | 33 (89) | 1.00 |
| Pedigrees | 37 (2; 1-15) |  |  |

Data are shown as median (interquartile range) and count (%). Pedigree data are shown as total number of pedigrees (median number of AFM per pedigree; range). Education level was assessed using the Verhage education system and dichotomised into low (level 1-5) and high (level 6-7) level of education. P-values were calculated using the Mann–Whitney U test for continuous variables and Fisher’s exact test for categorical variables. Abbreviations: AFM = asymptomatic family member; C9– = carriership of *C9orf72* with normal repeat length; C9+ = carriership of *C9orf72* repeat expansion.

### Physical examination and cognitive functioning do not differ between groups

No clinical signs of lower motor neuron involvement (e.g., muscle atrophy, weakness, fasciculations, hyporeflexia) were detected during physical examinations. No AFM reported symptoms of dysarthria, dysphagia, muscle weakness, hypertonia or new mobility issues. Signs that could be associated with mild upper motor neuron involvement were observed in both groups without significant group differences. Furthermore, there were no significant group differences in cognitive functioning on either ECAS or C-CAS. A detailed breakdown of these results is provided in Supplementary Table 2.

### Reduced periodic EEG power in *C9orf72* carriers

We investigated differences in EEG power between C9+ and C9-AFM. While the aperiodic component of the power spectrum (i.e., offset and exponent for the 2-48 Hz range) was not significantly different between the two groups, we found significant differences in the periodic component (Figure 1).

**Figure 1.**
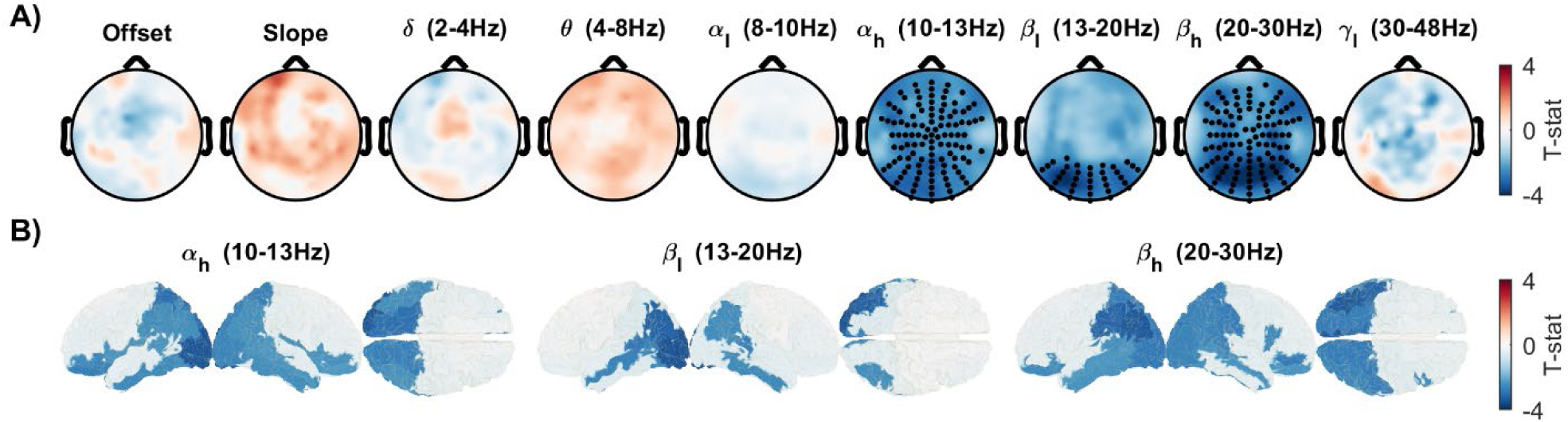
Differences in EEG power in asymptomatic *C9orf72* family members. **(A)** In sensor-space, no significant differences were found in the aperiodic power parameters (offset and exponent for the 2-48 Hz range), while periodic power in the α_l_, β_l_ and β_h_ frequency bands was lower in the C9+ group (bold black dots). **(B)** Source-space analysis corroborated these findings, showing periodic power differences in the same bands. These differences were localised primarily to posterior brain regions, encompassing the parietal, occipital, and temporal lobes. For the α_l_ and β_h_ bands, lower EEG power was also observed in the inferior frontal gyri. Abbreviations: AFM = asymptomatic family members; C9− = carriership of *C9orf72* with normal repeat length; C9+ = carriership of *C9orf72* repeat expansion.

In the sensor-space analysis, the C9+ cohort exhibited significantly lower EEG power compared to the C9-cohort in the α_l_, β_l_ and β_h_ frequency bands (Figure 1A). Source-space analysis corroborated these findings, localising the anatomical origins of this reduced power. For all three bands, the difference was primarily driven by posterior brain regions, encompassing the parietal, occipital, and temporal lobes. In the case of the α_l_ and β_h_ band, lower power was additionally observed in the inferior frontal gyri (Figure 1B).

The slope estimates of the aperiodic power in the higher-frequency rage (30-70 Hz), a proposed proxy of brain excitation/inhibition balance, showed no significant group differences in either sensor- or source-space (data not shown).

### Reduced EEG power is associated with *C9orf72* expression and cortical thickness

We next investigated the biological basis of the observed periodic EEG power reductions in α_l_, β_l_, and β_h_ by analysing their spatial patterns with normative *C9orf72* expression from the AHBA and with cortical thickness differences within the same cohort.

Spatial permutation testing revealed a significant negative relationship between the *C9orf72* expression pattern and the magnitude of power reduction in C9+ AFM (Figure 2A). Specifically, cortical regions with higher normative *C9orf72* expression were associated with a more pronounced decrease in EEG power in the C9+ group compared to the C9-group. These associations were found significant in all three tested frequency bands: α_l_ (P = 0.002), β_l_ (P = 0.029), and β_h_ (P < 0.001).

**Figure 2.**
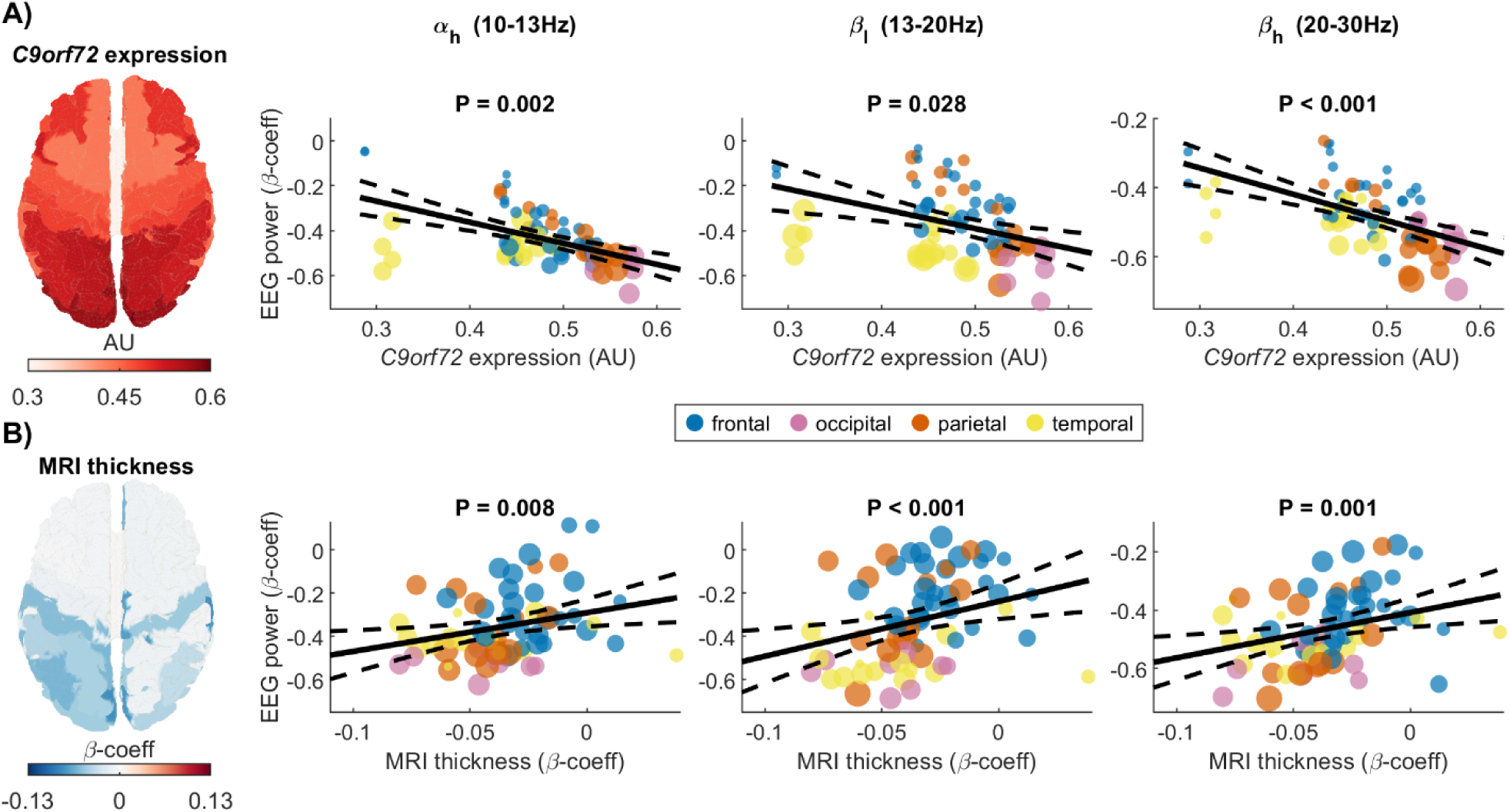
Relationship between reduced periodic EEG power, *C9orf72* expression, and cortical thickness. **(A)** Spatial permutation testing revealed a negative relationship between AHBA *C9orf72* expression and EEG power difference. The results show that regions with higher normative *C9orf72* expression exhibit a greater reduction (i.e., a more negative β coefficient) in α_l_, β_l_, and β_h_ EEG power in C9+ relative to C9-AFM. **(B)** The Spearman’s correlation analyses revealed a positive relationship between regional group differences in cortical MRI thickness and EEG power. These results show that regions with greater reduction in cortical thickness exhibit a greater reduction in α_l_, β_l_ and β_h_ EEG power in C9+ relative to C9-AFM. Only the regions showing significant difference (P_uncorrected_ < 0.05) in MRI thickness between the two groups were displayed on the brain plot. Abbreviations: AFM = asymptomatic family members; AHBA = Allen Human Brain Atlas; C9− = carriership of *C9orf72* with normal repeat length; C9+ = carriership of *C9orf72* repeat expansion.

Furthermore, Spearman’s correlation analyses demonstrated that this EEG power reduction was also significantly associated with structural MRI difference (Figure 2B). Brain regions with a greater reduction in cortical thickness also displayed a greater reduction in EEG power in the C9+ group compared to the C9-group. These associations were found significant in all three tested frequency bands: α_l_ (P = 0.008), β_l_ (P < 0.001), and β_h_ (P < 0.001).

### Reduced EEG power is related to visuospatial processing

To determine the functional relevance of the observed EEG power reductions, we next investigated their relationship with cognitive performance on specific C-CAS tasks that probe the functions of posterior brain regions: Visuoconstructive Immediate, Visuoconstructive Recall, and Body Representation.

Linear mixed-effects models revealed a significant interaction between C-CAS performance and *C9orf72* carriership in predicting periodic EEG power (Figure 3). The interactions showed that poorer performance on the Visuoconstructive Immediate task was significantly associated with lower EEG power in both the β_l_ and β_h_ bands in the C9+ cohort. Similarly, poorer performance on the Body Representation task was significantly associated with lower EEG power in the β_h_ band. These relationships were absent in the C9-cohort, and no significant associations were found for the Visuoconstructive Recall task in either group.

**Figure 3.**
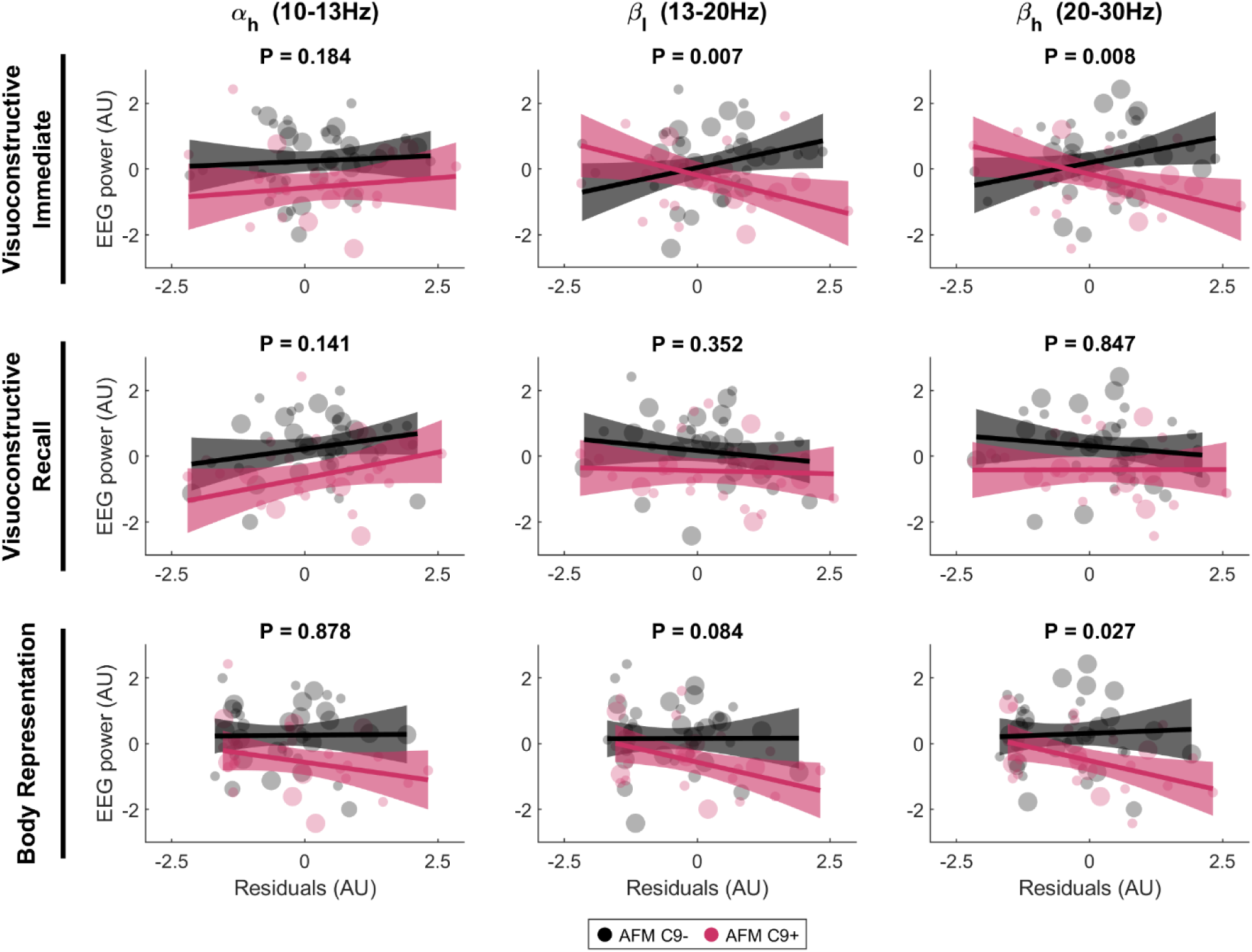
Relationship between reduced periodic EEG power and performance on visuospatial tasks. Linear mixed-effects models revealed a significant interaction between C-CAS performance and *C9orf72* carriership. Specifically, worse performance on Visuoconstructive Immediate and Body Representation tasks (indicated by higher residuals) was associated with lower β_l_ and β_h_ EEG power, an effect observed in C9+ AFM only. The reported P-values represent the interaction term (C-CAS × Group). For each participant, EEG power was estimated by averaging power values across the significant regions identified in the primary group analysis. Abbreviations: AFM = asymptomatic family members; C9− = carriership of *C9orf72* with normal repeat length; C9+ = carriership of *C9orf72* repeat expansion; C-CAS = Complementary Cognitive ALS Screen.

## Discussion

We demonstrate that resting-state EEG can detect functional alterations in the cerebral cortex of asymptomatic carriers of the pathological *C9orf72* RE. By comparing a cohort of carrier (C9+) and non-carrier (C9−) asymptomatic family members (AFM), we identified distinct EEG signatures of the *C9orf72* RE. Specifically, C9+ AFM exhibited significantly lower EEG power in the α and β (10–30 Hz) frequency range across the parietal, occipital, and temporal cortices. The magnitude of these electrophysiological alterations was correlated with reduced cortical MRI thickness within the same cohort and cortical *C9orf72* expression pattern from the Allen Human Brain Atlas (AHBA). These convergent findings suggest that the *C9orf72* RE may have an important involvement in the observed functional and structural differences, which may contribute to the deficits in visuospatial processing.

Oscillations in the α (8–12 Hz) and β (13–30 Hz) frequency bands are dominant rhythms in the resting brain, generated by the large-scale synchronisation of neural populations within corticocortical and thalamocortical networks.^24^ Our data demonstrate a reduction of these oscillations in the posterior brain regions of C9+ AFM, a finding that spatially correlates with decreased cortical thickness in the same cohort. A parsimonious explanation is that the observed loss of oscillatory power is a direct consequence of cortical atrophy, as a thinner cortex contains fewer neural generators. Interestingly, our longitudinal MRI study in an overlapping cohort revealed that C9+ AFM have a significantly faster rate of cortical thinning in the parietal regions over time.^5^ These findings suggest that the EEG changes we observe may be linked to an active process of structural change, a degenerative process occurring decades before clinical symptoms become evident, underscoring the potential importance of early therapeutic intervention.

At the molecular level, our findings may relate to the effects of the *C9orf72* RE. The spatial correlation between reduced EEG power and higher *C9orf72* expression suggests a connection between the functional brain changes and underlying molecular mechanisms, wherein both loss-of-function and gain-of-function are thought to create a vulnerable brain state.^25–27^ One downstream consequence of the *C9orf72* RE is evidenced to be a state of cortical hyperexcitability.^26–28^ Given that increased resting-state power in the α–β band is broadly understood to reflect a state of active functional inhibition,^29,30^ our data could suggest that these posterior brain areas are less inhibited in C9+ AFM. This interpretation, however, must be considered alongside our null finding for the aperiodic slope, a proposed proxy for the excitation/inhibition balance. While this measure has been validated in several studies, it has been primarily done in states of profound physiological change, such as in sleep or under anaesthetic agents.^31,32^ It may, therefore, lack the sensitivity to detect subtle or region-specific alterations in excitability that may be present in asymptomatic individuals. Our results, thus, only indicate that the overall aperiodic brain activity appears intact, and that further research is warranted to better delineate possible excitability changes.

The functional integrity of temporoparietal regions is crucial for executive control, providing transient inhibition via β-band burst to shape sensory processing and guide action.^30^ Our finding that reduced temporoparietal β-band power correlates with poorer performance on Visuoconstructive Immediate and Body Representation tasks supports this framework, as these tasks depend on spatial analysis and perceptual organisation functions of the posterior cortex.^33^ In addition, since this relationship was observed only in C9+ AFM, our data suggest a specific effect of the *C9orf72* RE on altered visuospatial processing. The absence of a relationship between EEG measures and the Visuoconstructive Recall task is intriguing and may reflect its reliance on both visuospatial and memory functions, which are likely to engage a broader network not fully captured in our analysis that focused only on posterior brain regions.

Our integration of functional, structural, gene expression, and cognitive data represents a crucial step towards a holistic understanding of the underlying processes in C9+ AFM. Specifically, by demonstrating a spatial correlation between reduced EEG power and cortical thinning, we show that structurally compromised regions are also functionally altered. Correlating EEG changes with *C9orf72* gene expression allows us to move from observation toward mechanistic inference. Furthermore, using data from cognitive screening, we provide evidence of a *C9orf72* RE effect on visuospatial processing. While this approach yielded novel insights, further understanding could be gained by exploring their relationship with additional data, such as excitatory/inhibitory receptor densities.

The identification of robust biomarkers that reflect deficits predictive of clinical conversion is an overarching aim with significant clinical implications. While our findings in resting-state EEG may represent a promising non-invasive, cost-effective, and scalable biomarker capable of serving multiple roles in future clinical trials, they should be interpreted in light of the cross-sectional design of our study. This design precludes any causal inferences regarding the trajectory of the observed changes. It remains unclear whether these EEG differences in asymptomatic carriers reflect neurodevelopmental alterations,^34^ compensatory mechanisms, or a neurodegenerative process. Disentangling these possibilities requires a longitudinal study that tracks these biomarkers over a longer period in relation to potential clinical conversion and other biomarkers.^35,36^ Furthermore, our study focused solely on cortical changes; hence, we can only draw conclusions regarding upper motor neuron involvement in the cerebral cortex, and we cannot infer involvement of subcortical regions or lower motor neurons. While subcortical regions are difficult to assess using EEG, future studies should incorporate measures that evaluate lower motor neurons.^37^

In conclusion, this study identifies reduced α–β oscillatory power as an electrophysiological signature of the *C9orf72* RE in asymptomatic carriers. We demonstrate that these functional brain differences correlate with patterns of structural brain differences and normative *C9orf72* expressions. Moreover, these neurophysiological changes were also linked to performance on visuospatial tasks, providing a potential basis for subclinical cognitive deficits. Together, these findings underscore the value of EEG as a non-invasive and cost-effective tool for identifying early functional brain alterations and support its utility in the development of risk biomarkers for *C9orf72* RE.

## Acknowledgements

We would like to express our gratitude to the staff of the Clinical Neurophysiology Department and the ALS Research Office at University Medical Centre Utrecht for their support. Our sincere thanks are also extended to all the study participants for their generous contribution of time to research.

## Data availability

The data that support the findings of this study are available from the corresponding author, upon reasonable request from qualified investigators, after permission from the appropriate regulatory authorities.

