## Supplementary Material for "Resting-state EEG oscillations are reduced in asymptomatic *C9orf72* repeat expansion carriers"

### Supplementary Table 1. Preprocessing outcomes and data quality assessments.

| | AFM C9- | AFM C9+ | P-value |
|---|---|---|---|
| Interpolated electrodes | 3 (1-6) | 3 (2-8) | 0.24 |
| ICA variance explained[a] (%) | 99.1 (98.6-99.4) | 99.0 (98.6-99.3) | 0.50 |
| Artifact ICs | 8 (6-12) | 10 (7-12) | 0.12 |
| Removed data (%) | 0.019 (0.005-0.066) | 0.023 (0.007-0.078) | 0.66 |
| Number of trials | 369 (361-375) | 370 (364-375) | 0.76 |

Data are shown as median (interquartile range). P-values were calculated using the Mann–Whitney U test. Abbreviations: AFM = asymptomatic family member; C9− = carriership of *C9orf72* with normal repeat length; C9+ = carriership of *C9orf72* repeat expansion; ICA = independent component analysis.

[a] ICA was performed after principal component analysis (PCA) reduction. A total of 70 PCA components were used to ensure that there were enough data points for each ICA component to be robustly estimated.

### Supplementary Table 2. Outcomes of physical examination and cognitive screening.

| | AFM C9- | AFM C9+ | P-value |
|---|---|---|---|
| **Physical examination**[a] | | | |
| Dysarthria | 0 (0) | 0 (0) | 1.00 |
| Impaired tongue movement | 0 (0) | 1 (3) | 1.00 |
| Sustained glabellar reflex | 1 (2) | 1 (3) | 0.85 |
| Jaw jerk reflex presence | 0 (0) | 2 (5) | 1.00 |
| Snout reflex presence | 8 (15) | 3 (8) | 0.25 |
| Palmomental reflex presence | 2 (4) | 6 (16) | 0.08 |
| Hypertonia arm muscles | 0 (0) | 0 (0) | 1.00 |
| Biceps tendon reflex<br>- Low-Normal<br>- Brisk<br>- Very brisk | 44 (83)<br>9 (17)<br>0 (0) | 28 (76)<br>9 (24)<br>0 (0) | 0.86 |
| Triceps tendon reflex<br>- Low-Normal<br>- Brisk<br>- Very brisk<br>- Missing data | 45 (85)<br>7 (13)<br>0 (0)<br>1 (2) | 29 (78)<br>7 (19)<br>0 (0)<br>1 (3) | 0.35 |
| Deltoid tendon reflex presence | 10 (19) | 10 (27) | 0.36 |
| Trapezoid tendon reflex presence | 8 (15) | 6 (16) | 0.63 |
| Pectoral tendon reflex presence | 5 (9) | 5 (14) | 0.40 |
| Hoffmann's reflex presence | 2 (4) | 3 (8) | 0.34 |
| Abdominal reflex absence | 4 (8) | 8 (22) | 0.07 |
| Hypertonia leg muscles | 0 (0) | 0 (0) | 1.00 |

| | | | |
|---|---|---|---|
| Knee jerk reflex<br>- Low-Normal<br>- Brisk<br>- Very brisk | 40 (75)<br>12 (23)<br>1 (2) | 29 (78)<br>6 (16)<br>2 (5) | 0.82 |
| Ankle jerk reflex<br>- Low-Normal<br>- Brisk<br>- Very brisk<br>- Missing data | 45 (86)<br>7 (13)<br>1 (2)<br>0 (0) | 29 (78)<br>5 (14)<br>1 (3)<br>2 (5) | 0.54 |
| Adductor reflex presence | 13 (25) | 9 (24) | 0.77 |
| Plantar reflex Babinski response | 0 (0) | 0 (0) | 1.00 |
| **Cognitive screening**[b,c] | | | |
| ECAS ALS specific | 87.0 (81.8-91.0) | 87.0 (82.0-91.0) | 0.92 |
| ECAS ALS nonspecific | 31.0 (29.0-33.0) | 31.0 (27.8-33.0) | 0.18 |
| ECAS Total | 118.0 (111.8-123.3) | 118.0 (111.5-122.0) | 0.66 |
| C-CAS Visuoconstructive Immediate | 0.16 (-0.37-0.85) | 0.33 (-0.52-1.02) | 0.75 |
| C-CAS Visuoconstructive Recall | 0.47 (-0.24-0.69) | 0.11 (-0.64-1.17) | 0.94 |
| C-CAS Body Representation | -0.31 (-1.29-0.11) | -0.23 (-1.33-0.77) | 0.50 |

The presence of the highest reflex or muscle tone, either on the left or right side, was used for each participant. Abbreviations: AFM = asymptomatic family member; ALS = amyotrophic lateral sclerosis; C9− = carriership of *C9orf72* with normal repeat length; C9+ = carriership of *C9orf72* repeat expansion; C-CAS = Complementary Cognitive ALS Screen; ECAS = Edinburgh Cognitive and Behavioural ALS Screen.

[a] Data are shown in count (%). P-values were calculated using ordinal regression or binomial logistic regression for dichotomous outcomes, with assessment outcome as the response variable and age, sex, and *C9orf72* RE carriership as predictors.

[b] ECAS data are raw scores shown as median (interquartile range). P-values were calculated using linear model analysis, with assessment outcome as the response variable and age, sex, education level, and *C9orf72* RE carriership as predictors. Lower scores indicate poorer performance.

[b] C-CAS data are model residuals shown as median (interquartile range). P-values were calculated using the Mann–Whitney U test, since the residuals were obtained using a normative model that already accounted for age, sex, and education. Higher residual indicate poorer performance.
